# Immunomodulatory Effects of Atractylodes Lancea in Healthy Volunteers with Dosage Prediction for Cholangiocarcinoma Therapy: a modelling approach

**DOI:** 10.1101/2023.05.08.23289655

**Authors:** Teerachat Sae-heng, Juntra Karbwang, Kesara Na-Bangchang

## Abstract

**Background:** A recent study on the immunomodulatory activity of *Atractylodes Lancea* (Thunb.) DC. (AL) in healthy Thai subjects revealed that a once daily dose of 1,000 mg AL administered for 21 days significantly inhibited the production of key pro-inflammatory cytokines, while stimulating the production of immune cells. There is however, no reported maximum tolerated dose (MTD) and suggested phase 2A dosage regimens in the literature.

**Objective:** This study aimed to evaluate the immunomodulatory effects *of Atractylodes Lancea* (Thunb.) DC. (AL) in healthy subjects, and to recommend optimal dose regimens for intrahepatic cholangiocarcinoma (iCCA) based on toxicity criteria.

**Methods:** A physiologically-based pharmacokinetic (PBPK) model, combined with the toxicological approach and the immunomodulatory effect, was used for a dose-finding. The safety and efficacy of each AL regimen were evaluated based on the previous study. At least, a daily OD dose of 1,000 mg AL significantly suppressed the production of all proinflammatory cytokines while significantly increasing the number of peripheral immune cells.

**Results:** The developed PBPK model well predicted clinical observed data. No significant differences in SII index values were found, but a difference in the lymphocyte-monocyte ratio was found on day 4. The dosage regimens for phase 2A are BID doses of 1,000 or 2,000 mg or OD doses of 2,000 mg. Preliminary results in phase 2A revealed that a once-daily dose of 2,000 mg had a significantly higher median overall survival, progression-free survival, disease control rate, and inhibition of increased tumor size without toxicities compared with a once-daily dose of 1,000 mg and standard supportive care.

**Conclusion:** A PBPK model, in conjunction with a toxicological approach, could assist in finding the potential dosage regimens for a clinical study, including herbal medicine.

## Introduction

Heterogeneous recruitment of various immune cells, i.e., CD8+, CD4+, natural killer (NK) cells, tumor-associated macrophages (TAM), cancer-associated fibroblasts (CAF), and myeloid-derived suppressor cells (MDSC) constitute an integral part of the tumor microenvironment (TME) of cholangiocarcinoma (CCA) [1]. With increasing knowledge of the TME, intrahepatic cholangiocarcinoma (iCCA) has been classified into four unique subtypes: immunological desert, immunogenic, myeloid, and mesenchymal [2]. This classification offers a new paradigm for iCCA therapy. Immune checkpoint inhibitors (ICIs) such as nivolumab and pembrolizumab are currently the research focus of iCCA therapy. PD1 and PD-L1 are programmed cell death proteins expressed in immune and cancer cells, respectively [3]. Anti-PD1 and anti-PD-L1 antibodies have been shown to improve the therapeutic efficacy in advanced-stage CCA patients with or without metastases [3]. In addition, nivolumab has been reported to enhance the clinical efficacy of the first-line treatment gemcitabine/cisplatin when combined in treatment-naive patients with advanced CCA [4]. Immunomodulators that regulate the TME thus present a novel treatment strategy for patients with advanced-stage CCA.

A recent study on the immunomodulatory activity of *Atractylodes Lancea* (Thunb.) DC. (AL) in healthy Thai subjects (atractylodin as the active ingredient) revealed that a once-daily dose of 1,000 mg AL administered for 21 days significantly inhibited the production of key pro-inflammatory cytokines while stimulating the production of immune cells [5]. Due to its potential immunomodulatory activity, AL could be a viable therapy for iCCA. A safe first-in-human (FIH) dose of AL has been proposed based on the maximum recommended starting dose (MRSD) determined in animal models [6]. There is, however, no reported maximum tolerated dose (MTD) and suggested dosage regimens for a phase 2A clinical study in the literature [5].

Physiologically based pharmacokinetic (PBPK) modeling is a technique accepted by the US FDA and EMA for drug submission that predicts optimal dose regimens for various disease therapies, including cancer therapeutics [7,8]. This approach avoids recruiting unnecessarily large numbers of research participants for clinical trials. This approach might be particularly advantageous for determining the optimal dose regimens of candidate drugs, notably anti-cancer drugs. The present study aimed to evaluate the immunomodulatory effects of AL in healthy subjects based on information from previous research [5,6] to predict FIH, MTD, and phase-2A dosing regimens of AL.

## Methods

### Data sources

The following data were obtained from a previous study in healthy Thai subjects: (i) plasma concentration-time profiles of atractylodin [6], the active component of AL, following a single dose of 1,000 mg AL (group 1) and once daily (OD) doses administered for 21 days (group 2), and (ii) parameters related to immunomodulatory activity, *i*.*e*., peripheral blood immune cells, and levels of pro-inflammatory cytokines following the OD of 1,000 mg AL or placebo administered for 21 days (group 2) [5]. This study was retrospectively registered on 17 October 2020 [Thai Clinical Trials Registry (TCTR: www.clinicaltrials.in.th) Number TCTR20201020001].The study was approved by the Ethics Committee of Thammasat University (No. 003/2564) and Sakhon Na-Kon Hospital (No. 049/2563). The study was conducted in accordance with Good Clinical Practice (GCP) guidelines and the Declaration of Helsinki.

### Pharmacokinetic analysis

The pharmacokinetic parameters of atractylodin were estimated using non-compartmental analysis (MonolixSuite Software, version 2021R1, Antony France, Lixoft, SAS, 2021). The parameters included area under the plasma concentration-time curve (AUC_0-inf_), maximum plasma concentration (C_max_), terminal half-life (t_1/2_), volume of distribution (V_z_/F), and clearance (CL/F).

### Statistical analysis

Peripheral blood immune cell data collected included in the analysis were: neutrophils, total lymphocytes, T cells, B cells, CD8+, CD4+, NK+ cells, CD4+/CD8+ ratio, lymphocyte-to-monocyte ratio (LMR), neutrophil-to-lymphocyte ratio (NLR), platelet-to-lymphocyte ratio (PLR), and systemic immune-inflammatory (SII) index. Circulating cytokines included IL-2, IL-4, IL-6, IL-10, TNF-α, and IFN-□. The distribution of all variables was assessed using the Shapiro-Wilk test. In addition, differences between the quantitative variables of the two groups were evaluated using the Pair-t test (normally distributed variables) and the Wilcoxon match-pair signed ranked test (non-normally distributed variables). The statistical significance level was set at α = 0·05.

### Model construction

A whole-PBPK model of atractylodin was developed based on our in-house and previously published data [9] using Simbiology®, a product of MATLAB 2018b (version 5.8.2, MathWorks, Natick, MA, USA). Model assumptions were: blood-flow restriction, rapid drug dissolution, absence of enterohepatic recirculation, and absence of stomach and large intestine absorption. The physicochemical and biochemical properties of atractylodin are summarized in **Table S1**.

### Model validation

The developed whole-PBPK model was validated against previously published clinical data. Absolute average-folding errors (AAFEs, equation 1), and virtual predictive checks (VPCs) were used to determine model validity (accepted AAFEs ≤ 2-fold) [10].

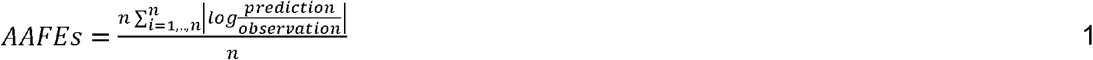

Where N is the number of parameters. Prediction, and observation are the predicted values and observed values, respectively.

### Sensitivity analysis

The sensitivity coefficient (equation 2) was computed to assess the uncertainty of the model parameters on atractylodin plasma-concentration profiles following OD dose of 1,000 mg AL for 21 days. A variation of 20% was applied to the fraction of unbound drug (f_u_), apparent permeability (P_app_), blood-to-plasma ratio (R_b:p_), solubility, and negative log of the acid dissociation constant (pKa).

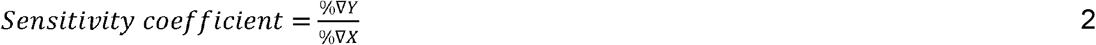

Where %∇*Y*, and %∇*X* are the percent changes of atractylodin concentration-time profiles, and model parameters, respectively.

### Prediction of MTD dosage regimens based on immunomodulatory activities

Plasma concentration-time profiles of atractylodin were initially simulated using the OD dose of 1,000 mg AL administered for 21 days, along with doses increased 2-, 4-, 6-, 8-, and 10-fold. The frequencies of administration were OD, twice-daily (BID), and four times-daily (QID). The maximum daily dose was 10,000 mg (10-fold) to minimize excessive dose administration, and drug bulk.

### Prediction of MTD dose regimens based on toxicity criteria

E_max_ model (equation 3) was applied to evaluate hematological toxicity of atractylodin on the surrogate peripheral blood mononuclear cells (PBMCs) (5) following various AL regimens, and was then compared with clinical data. The %E (reported as mean ± SD) was calculated to predict the maximal toxicity of atractylodin on PBMCs for each virtual population.

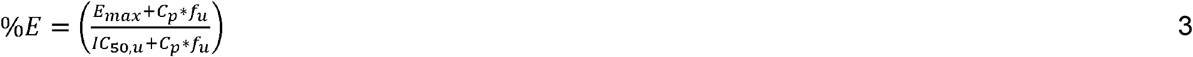

Where %E is the percentage of the toxicity of atractylodin on PBMCs. E_max_ is the maximum inhibitory effect (assumed to be 1 since the information is not available). C_p_ is atractylodin plasma concentration at time t. IC_50,u_ is half-maximum inhibitory effect of unbound atractylodin concentration (IC_50_=737 µmol/L) [6]. F_u_ is the fraction of unbound atractylodin in plasma.

Besides the risk of hematological toxicity, hepatotoxicity of atractylodin was assessed based on the ratios between C_max_ of unbound atractylodin in the liver (C_max, u, liver_) following the OD administration. The increases in the levels of liver enzymes, *i*.*e*., aspartate aminotransferase (AST), alanine transaminase (ALT), and alkaline phosphatase (ALP) were used as surrogate markers for the risk of hepatotoxicity associated with each AL regimen. A positive linear relationship between daily dose regimen (up to 10,000 mg), and liver enzyme levels was also assumed. The C_max, u, liver_ following the OD 1,000 mg regimen was considered non-hepatotoxic (no elevation of liver enzymes in any subject) (6).

### Virtual population simulation

%E, and the C_max, u, liver_ following various dose regimens of AL were simulated (Monte Carlo) with 100 virtual populations (18-60 years old, healthy subjects, average weight of 60 kg, and in fasting state). Since the overall survival (OS) in advanced stage CCA patients is 10 months [11], the risk of toxicity for this duration was predicted. For the purpose of predicting hematological and liver toxicity, AL treatment simulations were performed as 21-day courses.

## Results

The AUC_0-inf_, C_max_, V_z_/F, CL/F, and t_1/2_ for group 1 (day 1) and group 2 (days 1 and 21) are summarized in **Table 1**.

**Table 1.** Pharmacokinetic parameters following a single dose of 1,000 mg of AL administered (group 1), and daily doses of 1,000 mg of AL (group 2) administered for 21 days.

| Number | Parameter | Value |
| --- | --- | --- |
| Group 1 (Day 1) |  |  |
| 1 | AUC ($\mu\text{mol}\cdot\text{h/L}$) | 0.55 |
| 2 | $C_{\text{max}}$ ($\mu\text{mol/L}$) | 0.25 |
| 3 | $V_z/F$ (L/kg) | 11.33 |
| 4 | $CL/F$ (L/h/kg) | 5.01 |
| 5 | $T_{1/2}$ (h) | 1.63 |
| Group 2 (Day 1) |  |  |
| 6 | AUC ($\mu\text{mol}\cdot\text{h/L}$) | 0.64 |
| 7 | $C_{\text{max}}$ ($\mu\text{mol/L}$) | 0.28 |
| 8 | $V_z/F$ (L/kg) | 14.12 |
| 9 | $CL/F$ (L/h/kg) | 7.55 |
| 10 | $T_{1/2}$ (h) | 1.27 |
| Group 2 (Day 21) |  |  |
| 11 | AUC ($\mu\text{mol}\cdot\text{h/L}$) | 0.72 |
| 12 | $C_{\text{max}}$ ($\mu\text{mol/L}$) | 0.30 |
| 13 | $V_z/F$ (L/kg) | 13.41 |
| 14 | $CL/F$ (L/h/kg) | 8.04 |
| 15 | $T_{1/2}$ (h) | 1.14 |

AUC: Area Under Curve; $C_{\text{max}}$ : Peak plasma concentration; $CL/F$ : systemic clearance; $T_{1/2}$ : terminal half-life; $V_z/F$ : volume of distribution at steady state.

### Immunomodulatory effect of AL (atractylodin): Effects on circulating peripheral blood cells, immune cells, and pro-inflammatory cytokines

AL as an OD dose of 1,000 mg administered for 21 days markedly decreased the production of the pro-inflammatory cytokines IL-17A, IFN-□, TNF-α, IL-10, IL-6, IL-4, and IL-2 (**Table S2**). On the other hand, no significant decrease in the levels of these cytokines was observed in the placebo group (Table 2). In addition, AL significantly decreased the number of total lymphocytes and CD4+ cells (Table 3) while increasing B cells, T cells, CD8+ cells, NK cells, and the CD4+/CD8+ ratio (**Table S3**). In the placebo group, T cells, CD4+ cells, CD8+ cells, B cells, and NK cells were significantly reduced (Table 3). There was a significant change in LMR on day 4 in the AL-treated group and on day 22 in the placebo group (Table 3). The SII index values were comparable between the two groups (**Table S3**). The mean (±SD) of cytokine levels, immune cells, SII-index, and peripheral blood ratio on each day are shown in **Figures 1** and **2**.

**Table 2.** A statistical analysis of circulating cytokines following successive doses of 1,000 mg of AL administered for 21 days

| Treated group |  |  |  |  |
| --- | --- | --- | --- | --- |
| Cytokines | 0 hr. versus 24 hr. | 0 hr. versus 48 hr. | 0 hr. versus day 7 | 0 hr. versus day 14 |
| IL-2 | P=0.2121* | P=0.306* | P<0.0001* | P<0.001* |
| IL-4 | P=0.2524* | P=0.7948* | P=0.0002* | P=0.0007* |
| IL-6 | P=0.0637* | P=0.0826* | P=0.0353* | P=0.1303* |
| IL-10 | P=0.9853 (t=0.018, df=19) | P=0.87 (t=0.164, df=19) | P=0.0032 (t=3.378, df=19) | P=0.0566 (t=2.03, df=19) |
| IL-17A | P=0.0002* | P=0.0188* | P=0.0008 | P=0.0148 |
| TNF-α | P=0.063 (t=1.972, df=19) | P=0.529 (t=0.64, df=19) | P<0.0001 (t=4.94, df=19) | P=0.164 (t=1.448, df=19) |
| IFN-γ | P=0.044 (t=2.154, df=19) | P=0.0362* | P=0.1211* | P=0.0148 (t=2.68, df=19) |
| Untreated (Placebo) group |  |  |  |  |
| IL-2 | P=0.875* | P=0.625* | P=0.125* | P=0.125* |
| IL-4 | P=0.524 (t=0.72, df=3) | P=0.834 (t=0.23, df=3) | P=0.07 (t=2.76, df=3) | P=0.07 (t=2.76, df=3) |
| IL-6 | P=0.145 (t=1.95, df=3) | P=0.35 (t=1.13, df=3) | P=0.11 (t=2.22, df=3) | P=0.543 (t=0.69, df=3) |
| IL-10 | P=0.375* | P=0.25* | P=0.125* | P=0.625* |
| IL-17A | P=0.875* | P=0.5* | P=0.875* | P=0.125* |
| TNF-α | P=0.81 (t=0.26, df=3) | P=0.23 (t=1.51, df=3) | P=0.123 (t=2.59, df=2) | P=0.24 (t=1.47, df=3) |
| IFN-γ | P=0.88 (t=0.156, df=3) | P=0.28 (t=1.32, df=3) | P<0.0001 (t=96.85, df=3) | P=0.28 (t=1.32, df=3) |

\*Wilcoxon matched-pairs signed rank test; AL: Atractylodes Lancea (Thunb) D.C.; IL: interleukin; IFN- $\gamma$ : interferon-gamma; TNF-α : tumor-necrosis factor alpha

**Table 3.**
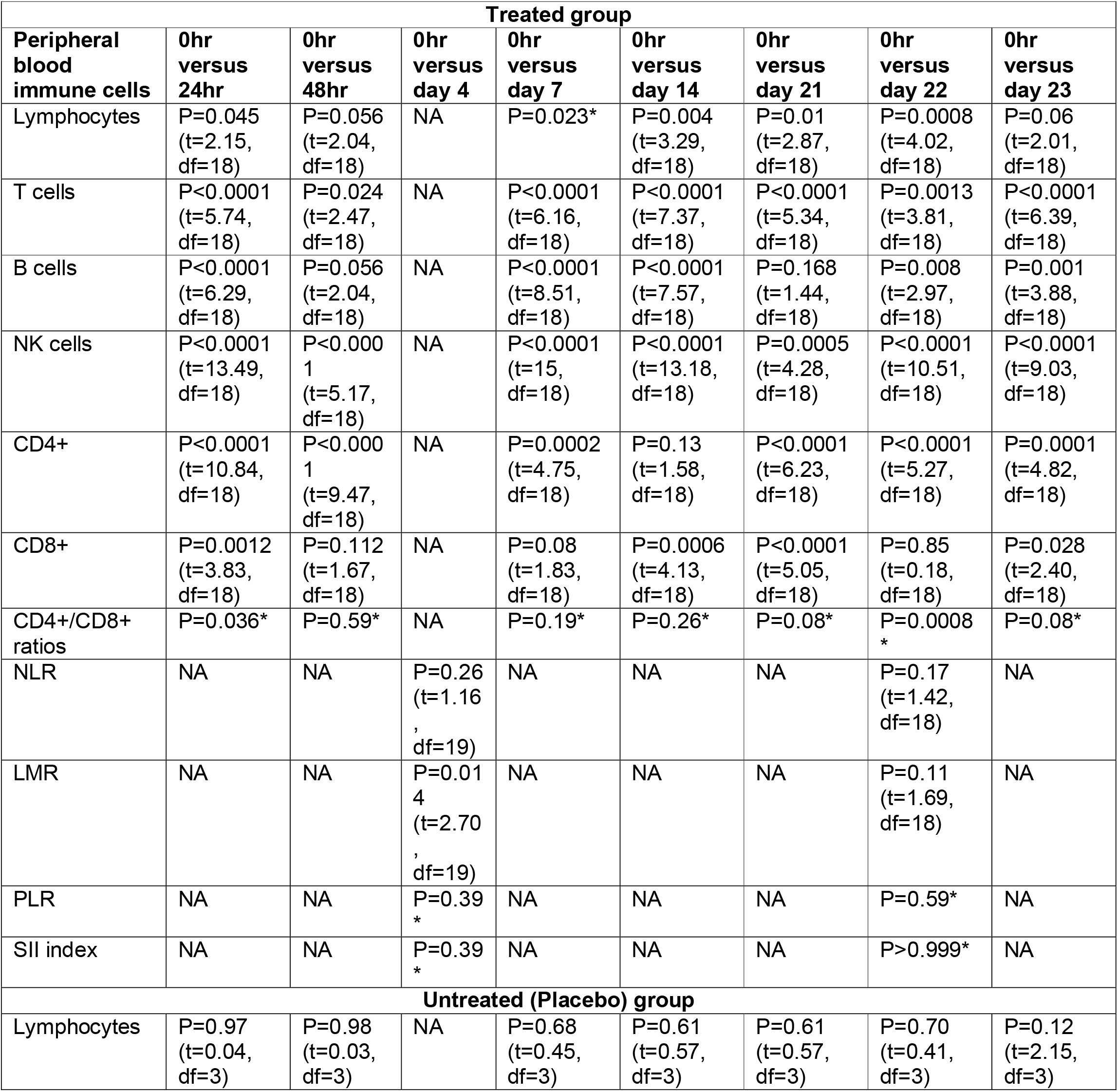
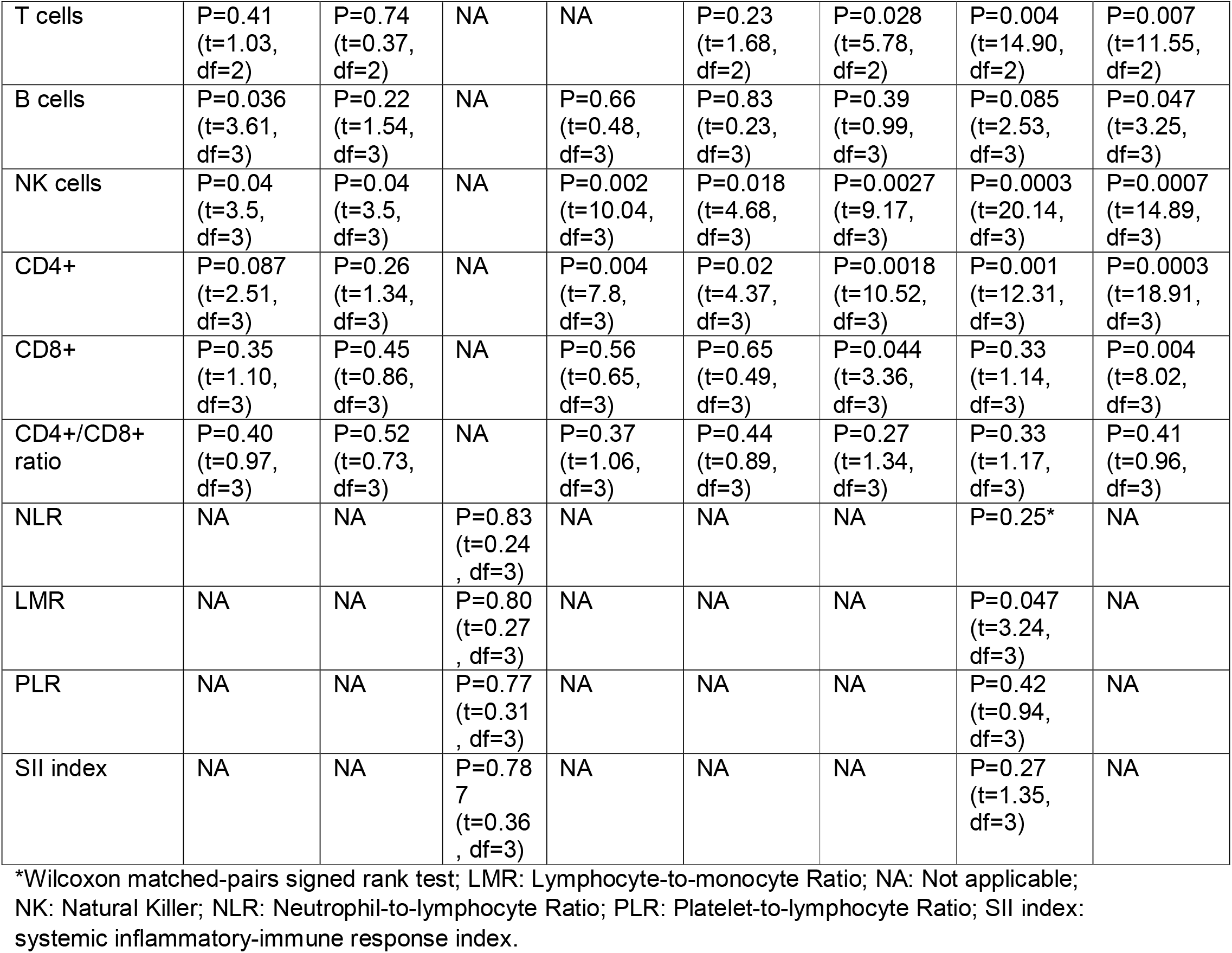
A statistical analysis of peripheral blood immune cells, SII index, NLR, LMR, PLR, and SII index following daily doses of 1,000 mg of AL administered for 21 days.

**Table 4.**
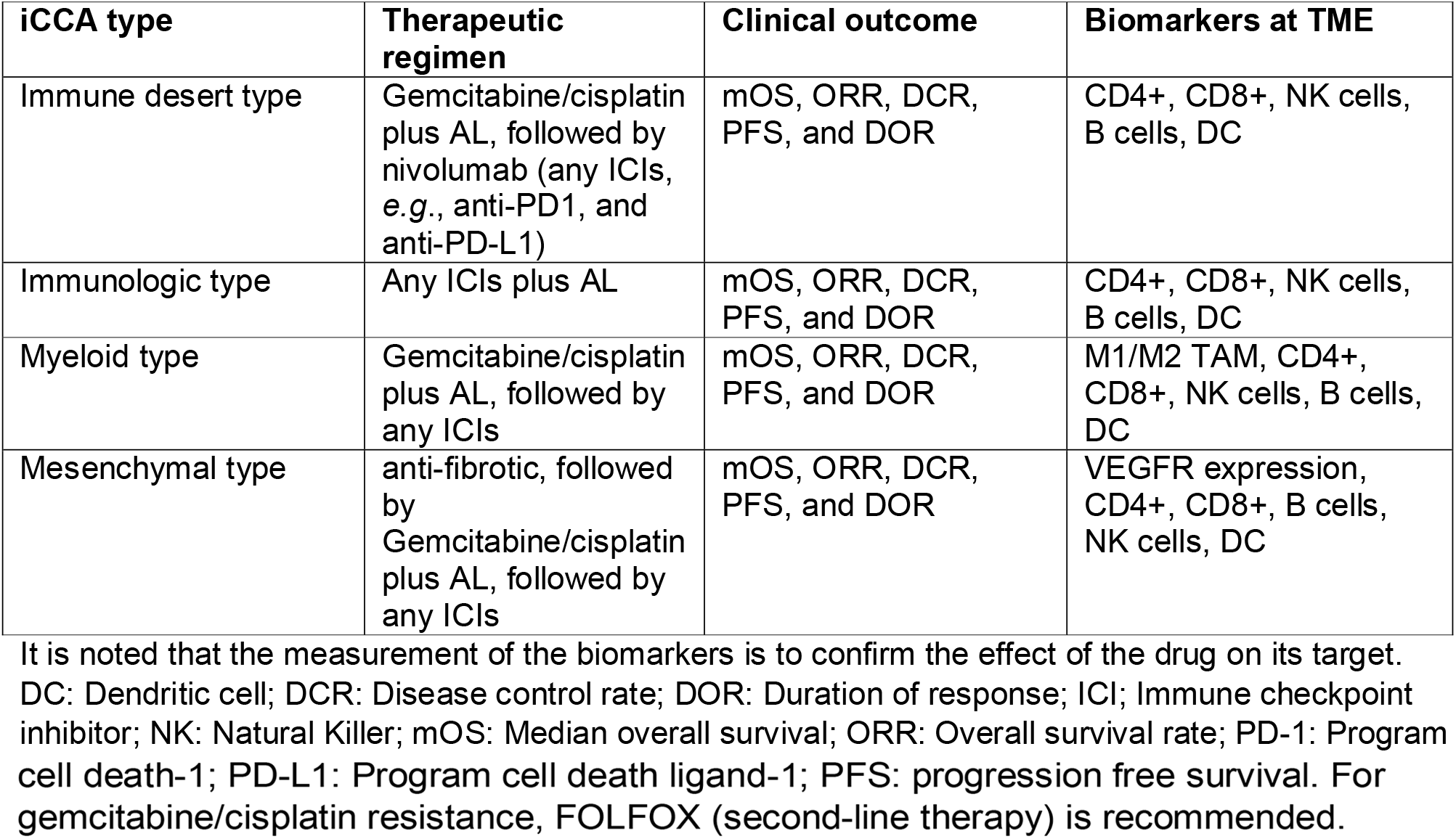
Recommended therapeutic regimens for intrahepatic cholangiocarcinoma (iCCA) based on immune tumor microenvironment (TME) classification.

| iCCA type | Therapeutic regimen | Clinical outcome | Biomarkers at TME |
| --- | --- | --- | --- |
| Immune desert type | Gemcitabine/cisplatin plus AL, followed by nivolumab (any ICIs, e.g., anti-PD1, and anti-PD-L1) | mOS, ORR, DCR, PFS, and DOR | CD4+, CD8+, NK cells, B cells, DC |
| Immunologic type | Any ICIs plus AL | mOS, ORR, DCR, PFS, and DOR | CD4+, CD8+, NK cells, B cells, DC |
| Myeloid type | Gemcitabine/cisplatin plus AL, followed by any ICIs | mOS, ORR, DCR, PFS, and DOR | M1/M2 TAM, CD4+, CD8+, NK cells, B cells, DC |
| Mesenchymal type | anti-fibrotic, followed by Gemcitabine/cisplatin plus AL, followed by any ICIs | mOS, ORR, DCR, PFS, and DOR | VEGFR expression, CD4+, CD8+, B cells, NK cells, DC |

It is noted that the measurement of the biomarkers is to confirm the effect of the drug on its target. DC: Dendritic cell; DCR: Disease control rate; DOR: Duration of response; ICI: Immune checkpoint inhibitor; NK: Natural Killer; mOS: Median overall survival; ORR: Overall survival rate; PD-1: Program
cell death-1; PD-L1: Program cell death ligand-1; PFS: progression free survival. For gemcitabine/cisplatin resistance, FOLFOX (second-line therapy) is recommended.

**Figure 1.**
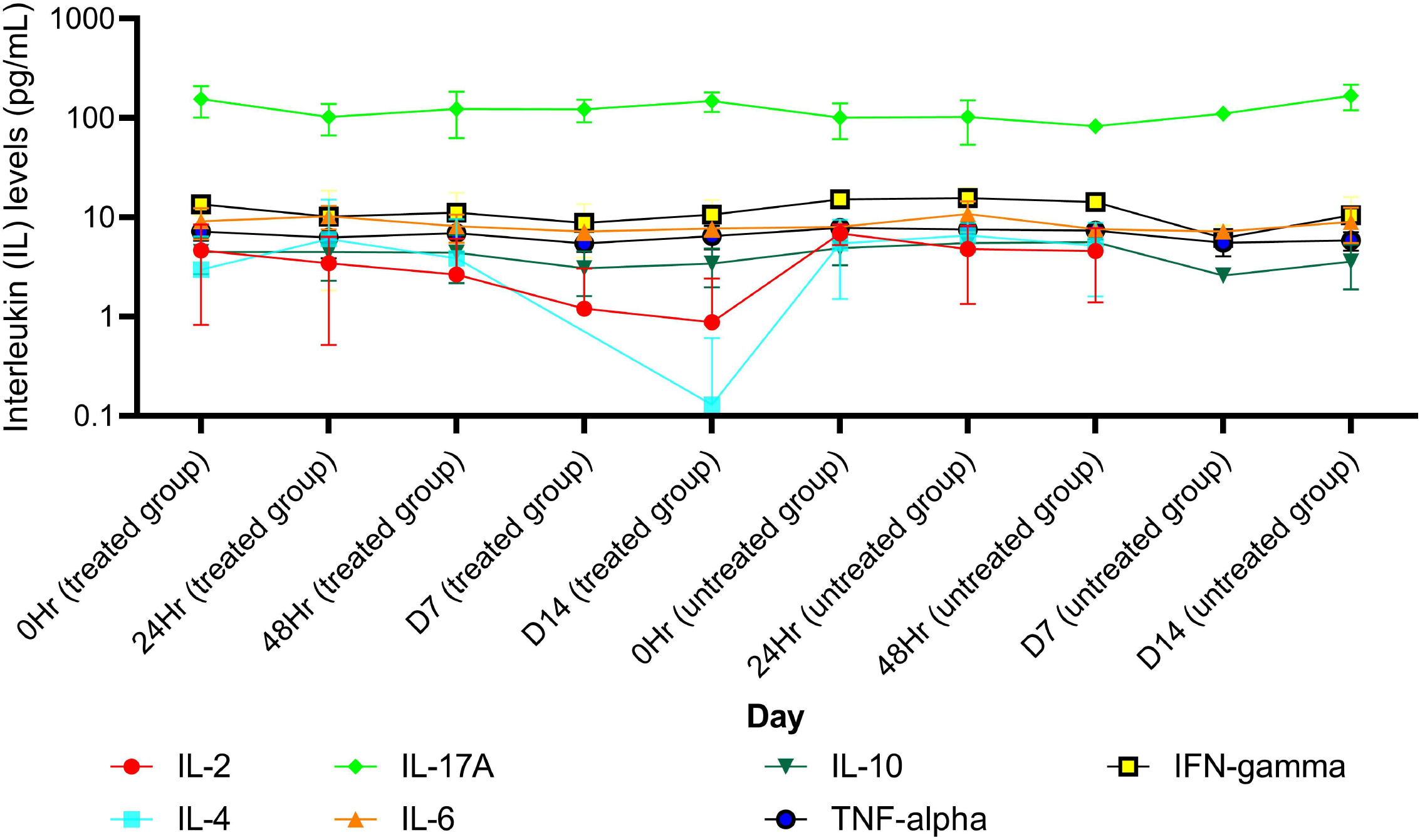
A comparison of different interleukins (IL) levels between treated group and untreated group for each day

**Figure 2.**
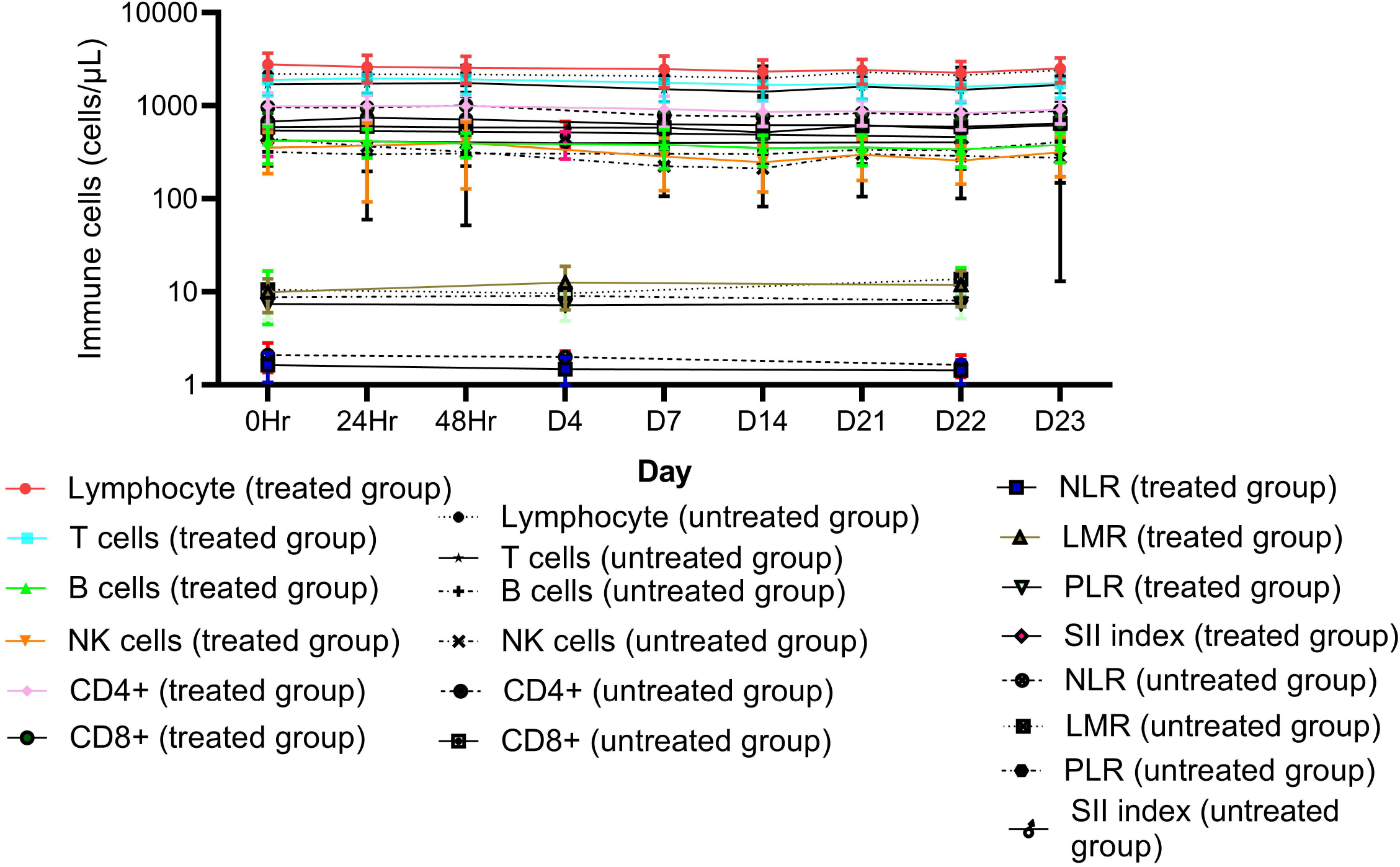
A comparison of different peripheral blood immune cells, peripheral blood index, and SII index between treated group and untreated group for each day

### Model validation and sensitivity analysis

The overall AAFEs was 1.17. AAFEs for AUC_0-inf_, C_max_, CL/F, V_z_/F, and t_1/2_ for each group are summarized in **Table S4**. VPCs (predicted versus clinical values) are shown in **Figures S1-S3**. The sensitivity coefficients for P_app_, R_b:p_, solubility, pK_a_, and f_u_ were +0.56, -0.90, +0.058, +0.14, and -0.18, respectively. A schematic workflow is shown in **Figure 3**.

**Figure 3.**
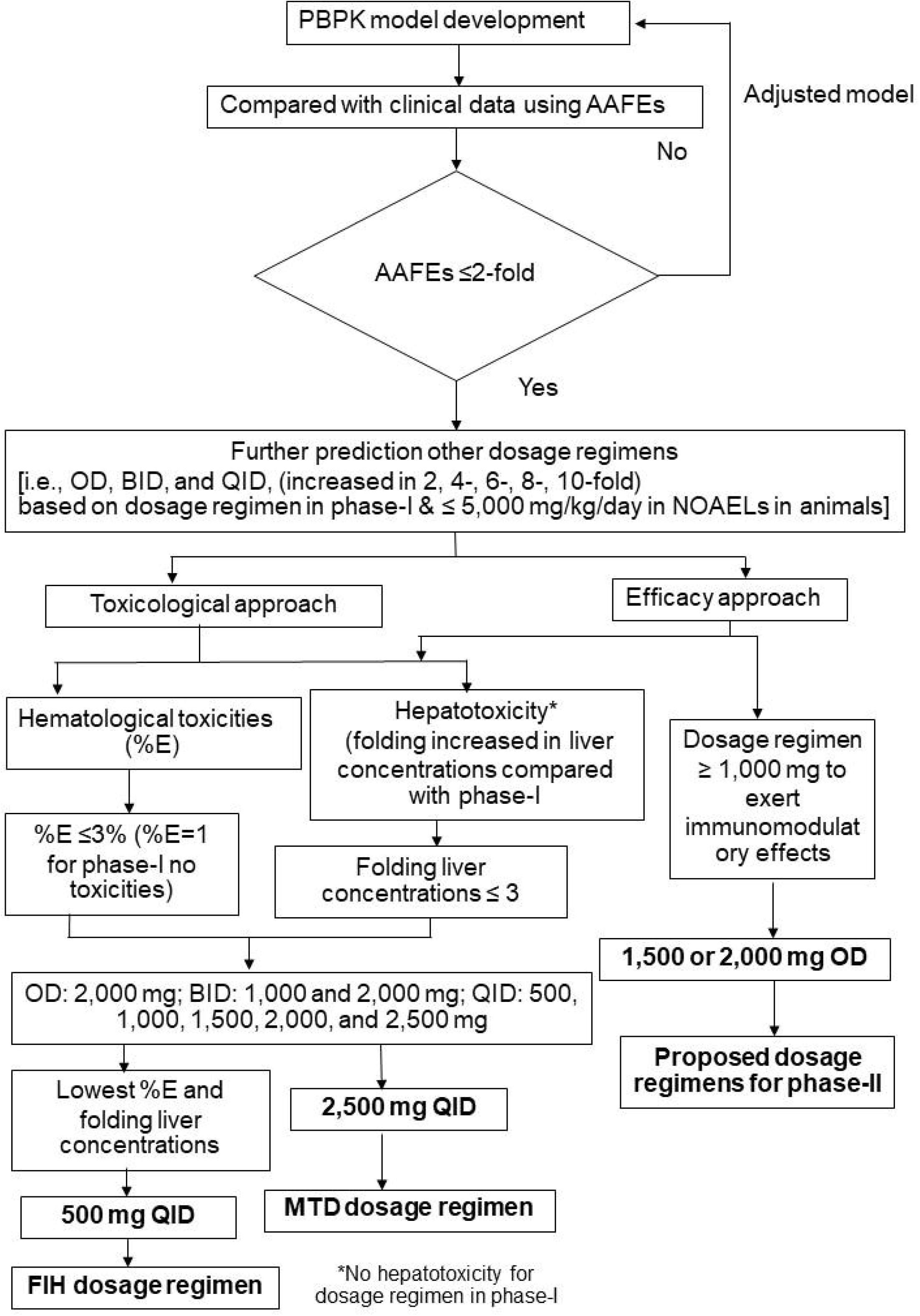
Schematic workflow criteria for dose selection

### Safety assessments of the simulated AL regimens

#### Hematological toxicities

The risks of hematological toxicity (%E) following an OD dose of 1,000 mg AL administered for 21 days and ten months were 1·06 ± 0·31% and 1·08±0·29%, respectively. When the dose was increased 2-fold (2,000 mg), 4-fold (4,000 mg), 6-fold (6,000 mg), 8-fold (8,000 mg), and 10-fold (10,000 mg), the risk increased to 2.24 ± 0.70%, 5.26±1.38%, 7.30 ±1.73%, 8.76 ±2.39, and 9.44 ±2.36 %, respectively. The %E following the BID dose of 1,000 mg AL (2,000 mg total) administered for ten months was 1.13 ±0·32%. When each dose of the BID regimen was increased to 2,000 mg (4,000 mg total), 3,000 mg (6,000 mg total), 4,000 mg (8,000 mg total), and 5,000 mg (10,000 mg total), the %E increased to 2.21±0.68%, 3.8 ±1.09%, 5.61 ±1.65%, and 6.89 ±1.5 %, respectively. The %E following the QID dose of 500 mg AL (2,000 mg total), 1,000 mg AL (4,000 mg total), 1,500 mg AL (6,000 mg total), 2,000 mg AL (8,000 mg total), and 2,500 mg AL (10,000 mg total) administered for ten months were 0.30 ±0.11%, 1.29 ±0.53%, 1.95 ±0.54%, 2.45 ±0.62%, and 3.27 ±0.68%, respectively.

#### Hepatotoxicity

The C_max, u, liver_ ratios of AL, when given an OD dose of 1,000 mg for both 21 days and ten months, increased 1.15-fold. When AL administration was extended to 10 months, increasing AL doses to 2,000, 4,000, 8,000, and 10,000 mg resulted in 1.40-, 3.73-, 5.40-, 6.50-, and 7.07-fold increases in liver enzyme levels, respectively. For the 10-month BID regimens, AL at 1,000, 2,000, 3,000, 4,000, and 5,000 mg resulted in 0·67-, 1.34-, 2·46-, 3.82-, and 4.74-fold increases in liver enzyme levels, respectively. For the 10-month QID regimens, AL at the doses of 500, 1,000, 1,500, 2,000, and 2,500 mg resulted in 0.17-, 0.71-, 1.07-, 1.40-, and 1.9-fold increases in liver enzyme levels, respectively.

## Discussion

The PBPK model developed here is found to be credible as the AAFEs were within 1.2-fold (≤2-fold), and none of sensitivity coefficients were higher than 1.

### FIH dose regimen

For the FIH dose regimens, the QID doses of 500 mg AL resulted in the lowest risks of hematological and liver toxicities compared with other regimens. Even with the higher total daily dose of 2,000 mg (human equivalent dose or HED = 2,400 mg) [6], which is about 2.5-fold lower than the observed non-observed adverse effect level (NOAEL) of 5,000 mg/kg observed in animals [6], the risks of hematological and liver toxicities remained low. The FIH dose, determined based on the toxicological criteria, appears safe and is lower than that used in the previous phase 1 study. A result from the phase 1 clinical trial suggests that this estimation of FIH, based on the information obtained from a pilot study and PBPK modeling, would provide a safe starting dose regimen for a drug candidate and avoid using an unnecessarily large number of research participants.

### The MTD regimens

The %E following the OD dose of 1,000 mg AL administered for 21 days was low (1%). This finding is consistent with the clinical data [6], supporting the validity of the Emax model for predicting the risk of hematological toxicity. The MTD dose regimen for AL was the QID dose of 2,500 mg AL given for up to 10 months, with a %E of ≤5%, resulting in 2-fold increases in the upper limit normal (ULN) of AST, and ALT levels, indicating the high safety profiles of this drug even with the high dosage administration. This risk is relatively low, and the side effects associated with AL are uncommon. Notably, this dose level is only about 50% of the observed NOAEL in the animal study [5]. However, this dosage regimen might not be appropriate for clinical use due to the high bulk at 10,000 mg daily.

The risk of hematological toxicity following the OD regimen of 10,000 mg AL was lower than 10%. It did not impact hematological cells but was more potent than the BID regimen (also 10,000 mg total).

Similarly, the risk of hepatotoxicity following the OD and BID regimens seemed to be increased, with AST and ALT levels increasing to 7-fold (grade-3) and 5-fold (grade-2) the ULN, respectively. Therefore, these two regimens are unsuitable for MTD dosage regimens due to the high risk of hepatotoxicity.

It has been reported that the conventional chemotherapy for iCCA (gemcitabine/cisplatin) is associated with grades 3-4 hematological toxicities, e.g., neutropenia (odds ratio (OR)=2.80, p<0.001), leukopenia (OR=2.98, p<0.001), and anemia (OR=2·96, p<0.001) [12]. A significant decrease in neutrophils was observed in 25% of the patients [13,14], of which 40% developed secondary infections [14]. In addition, gemcitabine/cisplatin was associated with liver toxicities [4, 13, 14], such as elevated levels of ALT (51.2%) and AST (53.5%) by >3-fold of ULN [4]. For second-line FOLFIRI-based chemotherapy (5-fluorouracil/irinotecan), 14.3% of the patients had increased ALT levels of grade 1 or 2 (>3-fold of ULN), and 85.7% had ascites [13].

### Recommended AL dose regimens for phase-2A clinical trial

A 1,500 or 2,000 mg AL QID dose is advised for phase-2A clinical trials in patients with advanced-stage CCA. The risks of hematological and liver toxicities following both regimens were lower than 3% and 1·5-fold (<3-fold of ULN). Notably, toxicities following most dose regimens of 1,500 mg AL are unlikely to occur since the surrogate indicators for toxicities were similar to those described in the previous study (<2-fold of ULN) [5]. It is noted that a dose of AL of at least 1,000 mg/day is required for immunomodulatory effects to be seen [5]. The BID (1,000 and 2,000 mg) and OD (2,000 mg) regimens are promising for phase-2A trials since the risks of toxicities were lower than QID regimens with higher compliance. Therefore, the OD dose of 1,000 and 2,000 mg was used in phase 2A clinical trial. Preliminary results revealed that a once-daily OD dose of 2,000 mg provided a significantly higher disease control rate (DCR), inhibition of increased tumor size, median overall survival (median OS), overall survival rate (OSR), progression-free survival (PFS), and progression-free survival rate (PFSR) in patients with unresectable or metastatic iCCA without toxicities compared with a once-daily OD dose of 1,000 mg.

### The roles of circulating peripheral cytokines and peripheral immune cells in iCCA therapy based on immunological classification

The increase in peripheral immune cells has been reported to be directly related to promoting immune cell recruitment in the TME [15]. The present study evaluated the effects of peripheral blood cells and pro-inflammatory cytokines on AL activity. Modulation (suppression or stimulation) of circulating peripheral immune cells, peripheral blood cells, and circulating cytokines may be surrogates for these cells at the TME and tumor sites. Recent research revealed an increase in OS (overall survival) for CCA patients with substantial infiltration of B lymphocytes [16]. AL has also been reported to facilitate the recruitment of immune cells to the TME by inhibiting the synthesis of matrix metalloproteinases (MMP-2 and MMP-9) [17], thus reducing fibrosis. AL also decreased the number of CD4+ cells. A high proportion of CD4+ circulating cells was associated with shorter recurrence-free survival (RFS) in CCA patients following surgery [18]. A decrease in LMR (≤4·17) was associated with favorable treatment outcomes in CCA [19]. AL raised LMR (figure 2) but differed minimally from the baseline. Any impact of AL on LMR was unlikely. However, information on the CD4+/CD8+ ratio in CCA is limited, although an increase in the CD4+/CD8+ ratio has been associated with an improved prognosis in hepatocellular carcinoma following transarterial chemoembolization (TACE) [20]. Besides peripheral immune cells, AL significantly reduced the production of all pro-inflammatory cytokines. Decreases in IL-4, IL-6, and IL-17A levels were associated with substantial improvements in CCA burden, OS, disease progression, and metastasis [21-25]. Inhibition of IL-4 production is crucial for the development of M2 TAMs and MDSC cells in the TME (26), as well as for the activation of mitogen-activated protein kinase (MAPK) [26]. A decrease in IL-4 receptor expression and increased tumor cell apoptosis was also observed [26]. IL-6 has been shown to inhibit CCA proliferation through the JAK-STAT3 (Janus kinase-signal transducer and activator of transcription-3) pathway and reduce the activity of M2 TAMs [27], thereby enhancing antitumor effects. It facilitates the recruitment of CD8+ and NK cells for IL-10 inhibition (CD94/NK group-2 member) [26]. IL-6 also suppresses the expression of lymphocyte activation gene-3 (LAG-3) and PD-1 (programmed cell death protein-1) on CD8+ cells in the TME [26], thus preventing T cell exhaustion. Suppression of IL-2 production improves the function of CD8+ and NK cells, increases the activity of regulatory T cells (Treg) at the IL-2 receptor, and suppresses CD8+ effector T cell function [28, 29]. A decrease in TNF-α and IFN-□ production might impair the functions of both NK and CD8+ cells. In the present study, however, there was only a modest change in the levels of both these cytokines in response to AL, compared with the baseline. The suppression of IL-10 expression by AL led to increased production of IFN-□ and TNF-α and promoted the function of CD8+ effector T cells.

Advances in the knowledge of TME and molecular analysis in recent years have facilitated the search for new iCCA treatments. By enhancing the effects of immunosurveillance, ICIs become a new possible therapeutic approach for iCCA therapy. As CD8+ function boosters, anti-PD1 (e.g., pembrolizumab and nivolumab) and anti-PD-L1 (e.g., atezolizumab) are effective against a variety of solid tumors, except iCCA [3]. Despite the expression of PD-L1 on tumor-associated immune cells, the depletion of T cells induced by drugs is unlikely to be adequate for iCCA therapy. However, nivolumab with gemcitabine/cisplatin has been reported to improve OS [4]. Gemcitabine may diminish the number of circulating immunosuppressive cells (i.e., MDSCs) and induce the expression of MHC-I (major histocompatibility complex) in cancer cells (increasing antigenicity) [30], which supports the function of CD8+ cells. The absence of T cells in CCA tumors attributes to the prevalence of immunological desert iCCA (I1: 48%) [2]. In contrast to other solid tumors, the treatment approach for iCCA involves transforming the tumor immunological type from cold to hot. The stromal cells and extracellular matrix (ECM) around the iCCA TME impede immune cell recruitment. Combining AL with gemcitabine/cisplatin, followed by an anti-PD1, is recommended for patients with advanced-stage iCCA. AL supports the recruitment of essential immune cells and facilitates dendritic cells (DC, MHC-I/II) to the tumor site by reducing fibrosis. On the other hand, Gemcitabine boosts the function of MHC-I/II by increasing the immunogenicity of iCCA, and anti-PD1 reinvigorates CD8+ cell function. During the treatment of immunogenic iCCA (I2: 9%) [2], immune-stimulating cells must outweigh the activity of immunosuppressive cells. The key characteristic of therapy for immunogenic iCCA is promoting the activity of immuno-stimulating cells to exceed that of immunosuppressive cells. A combination of nivolumab and AL is recommended. It has been reported that AL also supports the function of NK and CD8+ cytotoxic T cells. In addition, it inhibits the activity of Treg cells (through IL-2R inhibition) [28], which outweighs immuno-stimulating cell function. For myeloid iCCA (I3: 13%) [2], combination therapy of AL with any ICIs is recommended. Similarly, AL enhances the recruitment of antitumor immune cells, particularly CD8+ cells, to the tumor site. It also limits the recruitment of M2 TAMs by inhibiting the production of IL-4, and IL-10, thereby enhancing the functions of CD8+ and NK cells. ICIs also reverse the function of CD8+ cells in the TME. AL in combination with an anti-fibrotic agent, followed by gemcitabine/cisplatin, is suggested for mesenchymal iCCA (I4: 28%) [2] as it supports the reduction of tumor cell fibrosis through inhibition of MMP-2, and MMP-9 [17], thus augmenting the cytotoxic effects and facilitating the recruitment of CD4+, CD8+, and NK cells. For gemcitabine/cisplatin-resistant CCA, FOLFOX (folinic acid, 5-fluorouracil, and oxaliplatin), combined with ICIs, is recommended for increasing antigenicity [30].

The current study’s limitations include small sample size and the relatively low age of the virtual population (18-60 years in the placebo and AL-treated groups). In contrast, the average age of most patients with advanced-stage CCA is relatively older. The impact of the immunomodulatory effects of AL in patients with advanced-stage CCA may differ from healthy subjects. A phase-2A clinical trial with an adequate sample size is recommended to confirm the efficacy and safety of the proposed regimens. Since AL, specifically atractylodin, is an immunomodulator, the immune-modified response evaluation criteria for solid tumors (imRECIST) are suggested for evaluating tumor response endpoints due to the delayed impact of immunomodulators on tumor response.

In conclusion, the developed PBPK model accurately predicted the disposition of AL with AAFEs of 1·2-fold. Therefore, the suggested FIH and MTD regimens of AL are QID doses of 500 and 2,500 mg, respectively. In addition, the recommended dosage regimens for phase 2A are BID dose of 1,000 or 2,000 mg or OD dose of 2,000 mg. Notably, AL in conjunction with ICIs is recommended for each class of iCCA based on TME data in order to improve the clinical efficacy of AL.

## Data Availability

All data produced in the present study are available upon reasonable request to the authors.

## Acknowledgment

We thanks to dr. Marco Siccardi (Lab Drug Development, United Kingdom) for his assistant development of PBPK model. Also, dr. Rajith Rajoli (Department of molecular and clinical pharmacology, university of Liverpool) for his contribution for model development. In addition, we thank to dr. Tullayakorn Plengsuriyakarn, and dr. Nada Muhamad for their assistance of in-house model input parameters. We also appreciate the contributions for their comments from dr. Apichat Kaewdech, associate professor Teerha Piratvisuth, (Gastroenterology and hepatology unit, division of internal medicine, faculty of medicine, Prince of Songkla university).

## Authors’ contributions

TS: Conceptualization, data curation, validation, formal analysis, methodology, writing-original draft, interpretation. JK: Data curation, writing-review and editing. KN: Conceptualization, data curation, funding acquisition, supervision, project administration, writing-review and editing.

## Competing interests

The author(s) declare no competing interests.

## Consent for publication

Not applicable

## Data Availability

The data sets used and/or analysed during the current study are restricted due to the data contain potentially identifying or sensitive patient information. Contact information: (Ethical Committees).

## Ethic approval and consent to participate

This project has approval from the Human Research Ethics Committee of Thammasat University (Medicine) affiliated to Thammasat University (No. MTU-EC-00-3075/61). All participants signed written informed consent forms prior to enrollment.

## Funding information

This study was supported by Thammasat Postdoctoral Fellowship, Thammasat University Center of Excellence in Pharmacology and Molecular Biology of Malaria and Cholangiocarcinoma (No. 1/2556, dated 12 October 2013), and the National Research Council of Thailand (No. 45/2561, dated 10 September 2018). K.N. is supported by the National Research Council of Thailand under the Research Team Promotion grant (grant number NRCT 820/2563, dated 12 November 2020).

## Supporting information

Supplementary information is available at:

## Notes

### Competing Interest Statement

The authors have declared no competing interest.

### Clinical Trial

TCTR20201020001

