## Supplementary for "Immunomodulatory Effects of Atractylodes Lancea in Healthy Volunteers with Dosage Prediction for Cholangiocarcinoma Therapy: a modelling approach"

**Figure S1** A comparison of atractylodin plasma concentration-time profiles following a single dose of 1,000 mg of *Atractylodes Lancea* (Thunb) DC on day 1 between predicted values and clinical data.

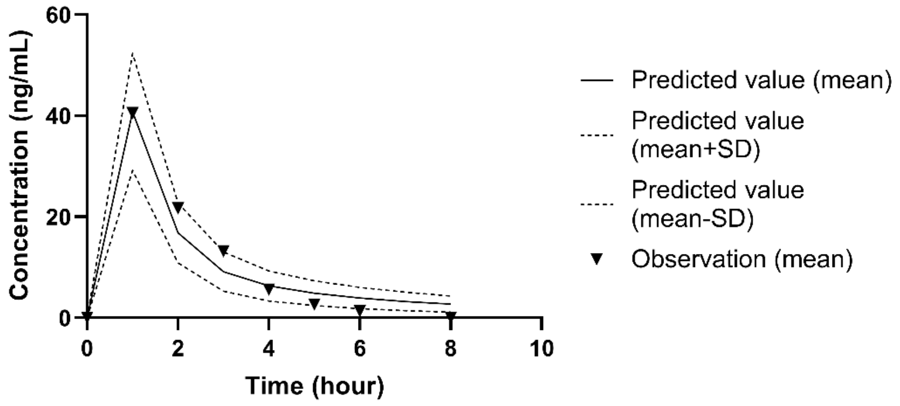


**Figure S2** A comparison of atractylodin plasma concentration-time profiles following multiple doses of 1,000 mg of *Atractylodes Lancea* (Thunb) DC on day 1 between predicted values and clinical data.

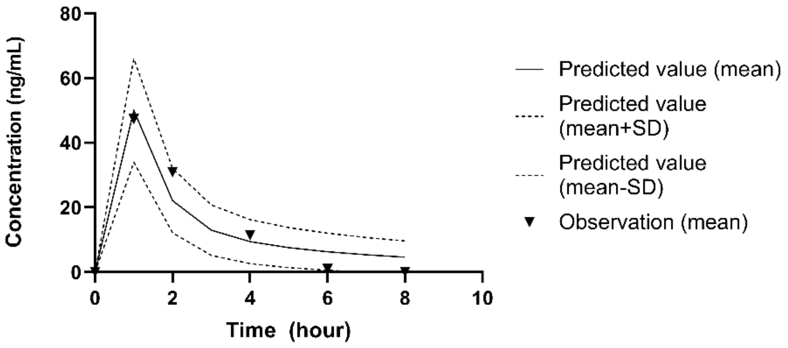


**Figure S3** A comparison of atractylodin plasma concentration-time profiles following multiple doses of 1,000 mg of *Atractylodes Lancea* (Thunb) DC on day 21 between predicted values and clinical data.

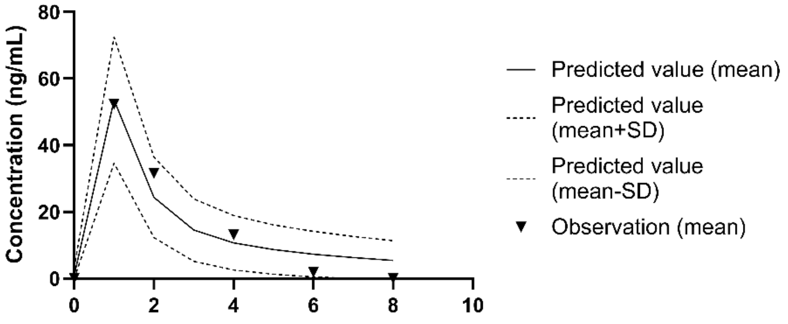


**Table 1.** Model parameters

| Number | Parameter | Value | Reference |
|---|---|---|---|

| | | | |
|---|---|---|---|
| **1** | Fraction of unbound drug ($f_u$) | 0.039 | In house |
| **2** | pKa | 9.63 | In house |
| **3** | Log D | 4.91 | Pub-chem |
| **4** | Blood-to-plasma partition ratio ($R_{bp}$) | 1.03 | In house |
| **5** | PSA | 13.1 | Pub-Chem |
| **6** | HBD | 1 | Pub-Chem |
| **7** | Absolute bioavailability ($F_{bio}$) | 0.8 | 1 |
| **8** | Clearance *in vivo* (L/h/kg) | 6.26 | 1 |
| **9** | Solubility (µg/ml) (PBS pH 6.8, SIF) | 0.11±0.003 | In house |

**Table 2**. Model validation

| **Number** | **Parameter** | **AAFEs** |
|---|---|---|
| | **All Group** | |
| **1** | AUC | 1.10 |
| **2** | $C_{max}$ | 1.06 |
| **3** | $V_z/F$ | 1.34 |
| **4** | CL/F | 1.13 |
| **5** | $T_{1/2}$ (h) | 1.22 |
| **AAFEs (Group 1; Day 1)** | 1.04 | |
| | **Group 1 (Day 1)** | |
| **6** | AUC | 1.02 |
| **7** | $C_{max}$ | 1.14 |
| **8** | $V_z/F$ | 1.02 |
| **9** | CL/F | 1.03 |
| **10** | $T_{1/2}$ | 1.00 |
| | **Group 2 (Day 1)** | |
| **11** | AUC | 1.19 |
| **12** | $C_{max}$ | 1.03 |
| **13** | $V_z/F$ | 1.49 |
| **14** | CL/F | 1.28 |
| **15** | $T_{1/2}$ | 1.27 |
| **AAFEs (Group 2; Day 1)** | 1.24 | |
| | **Group 2 (Day 21)** | |
| **16** | AUC | 1.11 |
| **17** | $C_{max}$ | 1.00 |
| **18** | $V_z/F$ | 1.60 |
| **19** | CL/F | 1.22 |
| **20** | $T_{1/2}$ | 1.42 |
| **AAFEs (Group 2; Day 21)** | 1.25 | |
| **AAFEs (All)** | 1.17 | |
